# The Accuracy of the Uganda National Tuberculosis and Leprosy Program algorithm and the World Health Organisation treatment decision algorithms for childhood tuberculosis: A retrospective analysis

**DOI:** 10.1101/2024.11.20.24317633

**Authors:** Peter J Kitonsa, Bernard Kikaire, Peter Wambi, Annet Nalutaaya, Jascent Nakafeero, Gertrude Nanyonga, Emma Kiconco, Deus Atwiine, Robert Castro, Ernest A Oumo, Hellen T Aanyu, Mary N Mudiope, Ezekiel Mupere, Moorine P Sekadde, Swomitra Mohanty, Adithya Cattamanchi, Eric Wobudeya, Devan Jaganath

## Abstract

Diagnosing childhood pulmonary tuberculosis (TB) is a challenge, and this led the Uganda National Tuberculosis and Leprosy Program (NTLP) to develop a clinical treatment decision algorithm (TDA) for children. However, there is limited data on its accuracy and how it compares to new World Health Organization (WHO) TB TDAs for children. This study aimed to evaluate and compare the accuracy of the 2017 Uganda NTLP algorithm with the 2022 WHO TDAs for TB among children in Kampala, Uganda.

We retrospectively assessed children <15 years old who underwent an evaluation for TB between November 2018 and November 2022. Children were classified as per National Institutes of Health (NIH) consensus definitions. We applied the 2017 Uganda NTLP and 2022 WHO algorithms (A with chest x-ray [CXR], B without CXR) to make a decision to treat for TB or not, and calculated accuracy in reference to Confirmed vs. Unlikely TB, as well as a microbiological and composite reference standard. We compared accuracy of the Uganda NTLP algorithm to the WHO TDAs among children <10 years old.

A total of 699 children were included in this analysis with 64% (451/699) under 5 years, 53% (373/669) were male, 12% (85/699) were Xpert Ultra positive, 11% (74/669) were HIV positive and 6% had severe acute malnutrition (SAM). The Uganda NTLP algorithm had a sensitivity of 97.9% (95% CI: 96.4-99.4) and specificity of 25.9% (95% CI: 21.2-30.7). If CXR was considered unavailable, sensitivity was 97.9% (95% CI: 96.4-99.4) and specificity 28.1% (95% CI: 23.2-33.0). Accuracy remained similar among high-risk children. In comparison, WHO TDAs had similar sensitivity to the Uganda NTLP, but algorithm A was more specific (32.2%, 95% CI: 26.9-37.5) and algorithm B was less specific (15.4%, 95% CI: 11.3-19.5).

Both the Uganda NTLP and WHO TDAs had high sensitivity but low specificity. The WHO TDAs had better specificity than the NTLP algorithm with CXR, and worse specificity without CXR. Further optimization of the algorithms is needed to improve specificity and reduce over-treatment of TB in children.

## INTRODUCTION

Childhood pulmonary tuberculosis (TB) presents a diagnostic challenge due to its non-specific presentation and difficulties in obtaining respiratory specimens because young children (who are most vulnerable) cannot expectorate. Additionally, sputum evaluation by smear and molecular testing is often negative due to the paucibacillary nature of childhood TB.^[1,2]^ Moreover, children often present at primary care facilities, which may lack the infrastructure for molecular testing or chest X-ray (CXR) to support TB evaluation and timely diagnosis. Consequently, there are large delays in TB treatment, which may explain why globally only 41% of the United Nations (UN) high-level meeting target of 3.5 million children on TB treatment was achieved.^[1]^

To reduce the delays in TB treatment initiation for children, treatment decision algorithms (TDAs) have been developed to guide providers on empiric treatment based on signs and symptoms. In Uganda, the National TB and Leprosy Program (NTLP) developed an algorithm that has been implemented since 2017.^[3]^ In 2022, the World Health Organization (WHO) conditionally recommended the use of TDAs for childhood TB, and presented an algorithm based on signs and symptoms as well as scores with CXR (Algorithm A) or without CXR (Algorithm B).^[4]^ However, there are limited data on the accuracy of these algorithms, and their use may result in over- or under-diagnosis of TB among children. Moreover, it is unclear if the new WHO algorithms should be utilized in Uganda or the current Uganda NTLP algorithm continued or updated.

To better define the role of these algorithms, we retrospectively analyzed four years of clinical data from children in Kampala, Uganda who underwent a standard TB evaluation and were classified according to National Institutes of Health (NIH) consensus definitions for childhood TB. We utilized this data to determine the accuracy of the Uganda NTLP algorithm, and also to compare its accuracy it to the WHO TDAs in same age group, and how accuracy varied based on access to Xpert and/or CXR.

## MATERIALS AND METHODS

### Study Population

De-identified data (accessed, 6^th^ December 2022) from children aged 0 - 14 years who underwent an evaluation for possible pulmonary TB between 28^th^ November 2018 and 30^th^ November 2022 was included for this analysis. The parent study recruited children from 28^th^ November 2018 to 19^th^ September 2024, situated at the Mulago National Referral Hospital Pediatric TB clinic within the Mulago National Referral Hospital (MNRH).^[5]^ The TB clinic recruited children from the MNRH pediatric wards, Mulago assessment center, other clinics and hospitals in and around Kampala (including Kampala Capital City Council (KCCA) clinics/health facilities and Infectious Diseases Institute (IDI) clinics), and referrals from upcountry health facilities. After evaluation, children returned to their referring facilities for further care. MNRH provides approximately 500,000 outpatient visits per year including from specialized clinics.^[5]^ The hospital’s immediate catchment area includes Kampala, Mukono and Wakiso districts with a population of approximately 1.8, 0.9 million and 3.4 million respectively^[6]^ with the main languages spoken being Luganda and English. The hospital is also a national center of excellence that attends to patients from the entire country.

Children were consecutively enrolled and included if they had cough at least 1 week and at least two of the following: unexplained fever >1 week, weight loss (>5% reduction compared to highest recorded weight in prior 3 months), poor appetite, poor weight gain/failure to thrive (weight-for-age or weight-for-height/length z-scores ≤-3), unexplained lethargy or reduced playfulness >1week, abnormal chest x-ray or history of TB contact. Children who were already on TB treatment, or history of TB disease in the last year, were excluded. Our analysis was restricted to children who met the Uganda intensified TB case finding (ICF) guide^[3]^ definition of presumptive TB applied at all entry points: any one of persistent cough ≥2 weeks or any duration if HIV positive: persistent fevers ≥2 weeks: poor weight gain in the last ≥1 month; TB contact, or swellings in the neck, armpit or groin.

### TB evaluation procedures

Participants completed an interview, which included a standardized questionnaire about TB symptoms and signs and the duration of each, along with questions about their sociodemographic information and potential TB risk factors. The clinical evaluation included physical examination, CXR, Tuberculin skin testing (TST), and HIV testing. Respiratory specimen collection included expectorated or induced sputum, gastric aspiration, or nasopharyngeal aspiration, and was sent for Gene Xpert MTB/RIF Ultra testing [Cepheid, Sunnyvale, USA] and solid or liquid mycobacterial culture. Xpert Ultra trace semi-quantitative results were considered positive. Additional details have been previously published.^[7]^

All individuals had their weight (in kg) and height/length (in meters) taken using a weighing scale and SECA-216 stadiometer (Seca Industries, Hamburg), respectively. All children had a follow up visit regardless of TB diagnosis at 2 months to assess ongoing or resolved signs and symptoms and any response to anti-TB treatment.

### Classification of TB status by Treatment Decision Algorithms

According to the NTLP algorithm^[3]^, if a child < 15 years old meets ICF criteria for presumptive TB, it is recommended to obtain a respiratory specimen for molecular testing with Xpert MTB/RIF Ultra. If MTB detected, they are treated for TB. If MTB is not detected, decision to treat was based on whether they met ≥2 of the following criteria if HIV negative OR ≥1 criteria if HIV positive; 1. 2 or more TB symptoms (persistent fever≥2 weeks, persistent cough≥2 weeks, poor weight gain ≥1 month), 2. positive history of TB contact (bacteriologically confirmed), 3. CXR suggestive of TB, and 4. any physical signs suggestive of TB. Physical signs suggestive of TB included: severe malnutrition, cervical, axilla or groin lymphadenopathy, features of pneumonia not responding to broad spectrum antibiotics.

The 2022 WHO TB treatment decision algorithms (TDAs) A and B were developed for children <10 years old, and so analysis was limited to this age group.^[8]^ The algorithms both recommend molecular testing for children with presumptive pulmonary TB. If negative or unavailable, a TB score is calculated that assigns points to TB signs and symptoms, with TB treatment decision when the total score ≥10. The algorithms differ in whether CXR is available (A) or is not (B), with corresponding different scores assigned to the TB features.

We analyzed TB treatment decisions for four scenarios: (1) when Xpert and CXR is available; (2) when only CXR is available; (3) when only Xpert is available; and (4) when neither Xpert nor CXR is available. The Uganda NTLP algorithm was also evaluated for all the four scenarios, and WHO algorithm A for (1) and (2), and algorithm B for (3) and (4).

### Reference Standard

Children were classified as Confirmed TB (positive Xpert or culture), Unconfirmed TB (negative microbiological testing but consideration of other signs and symptoms of TB and response to TB treatment) or Unlikely TB (not meeting other criteria) using the NIH consensus definitions.^[9]^ They were unclassifiable if there was insufficient data to determine TB status. Two pediatric TB experts classified each participant independently and blinded to the TDA calculation, with a third reviewer as a tiebreaker if needed. We used the following reference standards:

1. **Confirmed TB versus Unlikely TB**. This excludes children with Unconfirmed TB.
2. **Composite reference standard (CRS)**. Confirmed or Unconfirmed are defined as TB, and Unlikely as not having TB.
3. **Microbiologic reference standard (MRS)**. Confirmed TB was defined as TB, and Unconfirmed or Unlikely TB as not having TB.

### Statistical methods

Descriptive statistics were computed to describe relevant characteristics, and categorical variables were summarized using frequencies and proportions and all continuous variables were summarized using medians and interquartile range (IQR). Our primary objective was to determine the accuracy of the algorithms. Utilizing each reference standard above, we calculated the sensitivity and specificity of the Uganda NTLP algorithm (children <15years) and the WHO TDAs (children <10 years) for the overall and for key subgroups of children <5 years old, with SAM, or living with HIV.

We further compared the accuracy of the Uganda NTLP algorithm with that of the WHO TDAs, among children <10 years. We compared the sensitivity and specificity of each algorithm using McNemar’s test, with significance defined as p-value < 0.05.

Stata version 13.0 (Statacorp, College Station, USA) was used for this analysis.^[10]^

### Ethical Considerations

The study was approved by the Ethics Review Committee of the Makerere University School of Medicine Research and Ethics Committee as well as the Mulago Hospital Research and Ethics Committee, Kampala, Uganda (IRB number 12884), and the institutional review board at the University of California, San Francisco (UCSF), United States. All children 8 to <15 years provided informed written assent and parental/guardian written consent where as children <8 years, only parental/guardian informed written consent was obtained for all study activities.

## RESULTS

### Participant characteristics and TB

Between November 2018 to November 2022, there was a total of 875 children with TB symptoms of whom 867 met the intensified TB case finding (ICF) criteria, and of whom 842 were enrolled into the study (Figure 1). TB status was unclassifiable for 143 participants and 25 did not consent; and were excluded. We thus analyzed data from a total of 699 children, of whom 98 (14.1%) had Confirmed TB, 361 (51.6%) had Unconfirmed TB, and 240 (34.3%) had Unlikely TB.

**Figure 1:**
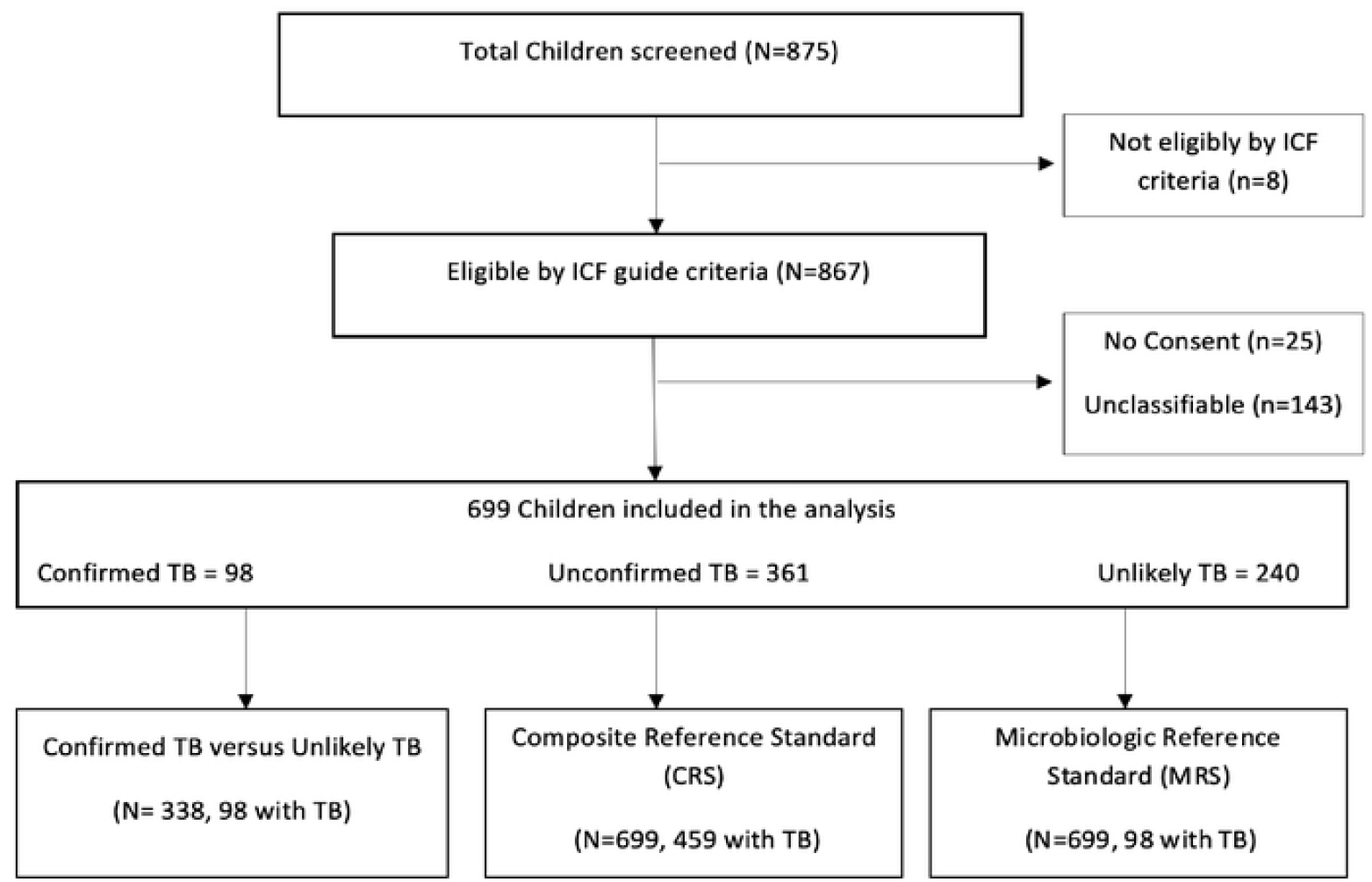
Study cohort flowchart.

The participants’ median age was 3 years (Interquartile Range, IQR: 1-6), 53% (373/699) were male, 64% (451/699) were aged <5 years, 92% (642/699) were aged <10 years, 57% (399/699) had history of TB contact, and 6% (42/699) had severe acute malnutrition (SAM). There were 74 (11%) children with HIV (CHIV), and 12% (85/699) of children had a positive Xpert result (Table 1).

**Table 1:** Characteristics of enrolled children with presumptive pulmonary tuberculosis in Kampala, Uganda (n=699)

| Characteristic | n (%) |
| --- | --- |
| Age Category |  |
| < 1 year | 80 (11.4%) |
| 1 - < 5 years | 371 (53.1%) |
| 5 - < 10 years | 191 (27.3%) |
| 10 - < 15 years | 57 (8.2%) |
| Male sex | 373 (53.4%) |
| Has fever<sup>1</sup> | 560 (80.5%) |
| Has lymphadenopathy<sup>1</sup> | 332 (47.6%) |
| Has weight loss | 271 (38.8%) |
| Failed to gain weight<sup>1</sup> | 584 (84.0%) |
| Has TB contact | 399 (57.1%) |
| HIV Positive<sup>1</sup> | 74 (11.1%) |
| Severe acute malnutrition | 42 (6.0%) |
| Chest X-ray interpretation<sup>1</sup> |  |
| Normal | 387 (61.5%) |
| Abnormal - Equivocal | 129 (20.5%) |
| Abnormal - Likely TB | 113 (18.0%) |
| Xpert MTB/RIF Ultra Positive | 85 (12.2%) |

1. 3 missing fever, 1 missing lymphadenopathy, 4 missing Failure to gain weight, 30 missing HIV status, and 70 missing chest-xray.

### Diagnostic accuracy of the Uganda NTLP Algorithm

When CXR was available, the overall sensitivity was 97.9% (95% CI: 96.4-99.4) among Confirmed vs. Unlikely TB, 90.7% (95% CI: 88.5-92.9) with the CRS, and 97.9% (95% CI: 96.4-99.4) with MRS. Specificity was 25.9% (95% CI: 21.2-30.7) in reference to Confirmed vs. Unlikely TB and the CRS, and 17.1% (95% CI: 14.3-20.0) with the MRS. (Table 2)

**Table 2:** Diagnostic accuracy of the 2017 Uganda national childhood TB diagnosis algorithm (NTLP) and the 2022 WHO TB diagnosis algorithms A & B under three reference standards with or without chest x-ray (CXR). Xpert results were incorporated into the TOA classification

| **Confirmed TB versus Unlikely TB** |  |  |
| --- | --- | --- |
| **Algorithm<sup>1,2</sup>** | **Sensitivity (95% CI)** | **Specificity (95% CI)** |
| **With CXR** |  |  |
| Uganda NTLP | 97.9 (96.4, 99.4) | 25.9 (21.2, 30.7) |
| WHO Algorithm A | 96.3 (94.2, 98.5) | 32.2 (26.9, 37.5) |
| **Without CXR** |  |  |
| Uganda NTLP | 97.9 (96.4, 99.4) | 28.1 (23.2, 33.0) |
| WHO Algorithm B | 97.5 (95.8, 99.3) | 15.4 (11.3, 19.5) |
| **Composite Reference Standard (CRS)** |  |  |
| **Algorithm** | **Sensitivity (95% CI)** | **Specificity (95% CI)** |
| **With CXR** |  |  |
| Uganda NTLP | 90.7 (88.5, 92.9) | 25.9 (22.6, 29.2) |
| WHO Algorithm A | 89.3 (86.8, 91.7) | 32.2 (28.5, 35.9) |
| **Without CXR** |  |  |
| Uganda NTLP | 88.4 (86.0, 90.9) | 28.1 (24.7, 31.5) |
| WHO Algorithm B | 94.2 (92.4, 96.1) | 15.4 (12.5, 18.2) |
| **Microbiologic Reference Standard (MRS)** |  |  |
| **Algorithm** | **Sensitivity (95% CI)** | **Specificity (95% CI)** |
| **With CXR** |  |  |
| Uganda NTLP | 97.9 (96.8, 99.0) | 17.1 (14.3, 20.0) |
| WHO Algorithm A | 96.3 (94.9, 97.8) | 20.4 (17.3, 23.5) |
| **Without CXR** |  |  |
| Uganda NTLP | 97.9 (96.8, 99.0) | 19.7 (16.7, 22.8) |
| WHO Algorithm B | 97.5 (96.3, 98.8) | 10.1 (7.7, 12.4) |

CXR- chest x-ray
WHO Algorithm A-settings with chest X-ray.
WHO Algorithm B-settings without chest X-ray
1. Algorithms were applied to the age group they were indicated for: the NTLP for children <15 years, and <10 years for the WHO TDAs
2. Xpert Ultra results included in the algorithms

When CXR was unavailable, the sensitivity remained the same in reference to Confirmed vs. Unlikely TB and the MRS, and reduced modestly with the CRS from 90.7% to 88.4%. In contrast, there were small increases in specificity and ranged from 19.7% to 28.1%. (Table 2)

### Accuracy of the Uganda NTLP algorithm by risk group

We evaluated the accuracy of the NTLP algorithm among children less than 5 years, CHIV, and those with SAM (Table 3). For each reference standard, the sensitivity and specificity remained similar to the overall estimate with overlapping confidence intervals. Similar to the overall estimate, the specificity slightly increased without CXR.

**Table 3:** Accuracy of the 2017 Uganda national childhood TB diagnosis algorithm (NTLP) by risk group (Age < 5 years, SAM, and HIV) under three reference standards with or without chest x-ray (CXR) for children <15 years old.

| **Confirmed TB versus Unlikely TB** |  |  |
| --- | --- | --- |
| Group | Sensitivity (95% CI) | Specificity (95% CI) |
| **Age < 5 years** |  |  |
| With CXR | 98.3 (96.5, 100.1) | 29.4 (23.0, 35.7) |
| Without CXR | 98.3 (96.5, 100.1) | 32.3 (25.8, 38.8) |
| **SAM** |  |  |
| With CXR | 97.6 (95.9, 99.3) | 25.7 (20.7, 30.7) |
| Without CXR | 97.6 (95.9, 99.3) | 28.1 (22.9, 33.2) |
| **HIV** |  |  |
| With CXR | 97.5 (95.7, 99.3) | 25.7 (20.6, 30.7) |
| Without CXR | 97.5 (95.7, 99.3) | 28.1 (22.9, 33.2) |
| **Composite Reference Standard (CRS)** |  |  |
| Group | Sensitivity (95% CI) | Specificity (95% CI) |
| **Age < 5 years** |  |  |
| With CXR | 89.9 (87.0, 92.7) | 29.4 (25.1, 33.7) |
| Without CXR | 88.2 (85.1, 91.2) | 32.3 (27.9, 36.7) |
| **SAM** |  |  |
| With CXR | 90.6 (88.3, 92.9) | 25.7 (22.2, 29.1) |
| Without CXR | 88.9 (86.5, 91.4) | 28.1 (24.5, 31.6) |
| **HIV** |  |  |
| With CXR | 90.5 (88.2, 92.8) | 25.7 (22.2, 29.1) |
| Without CXR | 88.8 (86.3, 91.3) | 28.1 (24.5, 31.6) |
| **Microbiologic Reference Standard (MRS)** |  |  |
| Group | Sensitivity (95% CI) | Specificity (95% CI) |
| **Age < 5 years** |  |  |
| With CXR | 98.3 (97.1, 99.5) | 18.5 (14.8, 22.2) |
| Without CXR | 98.3 (97.1, 99.5) | 20.9 (17.1, 24.8) |
| **SAM** |  |  |
| With CXR | 97.9 (96.8, 99.0) | 19.7 (16.7, 22.8) |
| Without CXR | 97.6 (96.4, 98.8) | 19.1 (16.0, 22.2) |
| **HIV** |  |  |
| With CXR | 97.5 (96.2, 98.7) | 16.8 (13.9, 19.8) |
| Without CXR | 97.5 (96.2, 98.7) | 19.1 (16.0, 22.2) |

### Accuracy of the WHO algorithms A and B

The accuracy of the WHO TDAs (children <10 years) is shown in Table 2. Similar to the NTLP, sensitivity was high but specificity was low across reference standards. However, in contrast, specificity varied depending on availability of CXR, with higher specificity when CXR was available for WHO algorithm A.

### Comparison of the diagnostic accuracy of the 2017 Uganda NTLP algorithm and the 2022 WHO childhood TB diagnosis algorithms

Among children <10 years, the accuracy of the Uganda NTLP algorithm and the WHO TDAs varied depending on the availability of Xpert and/or CXR (Table 4). We found that across the scenarios, the sensitivity between the Uganda NTLP and WHO TDAs remained similar, except WHO TDA-B had higher sensitivity compared to Uganda NTLP algorithm when CXR was not available against the CRS only (94.2% vs. 87.5% without CXR or Xpert, difference of −7.0, [95% CI: −10.7, −3.2], p <0.01). Specificity depended on the availability of CXR; if CXR was available, WHO TDA-A was more specific than the Uganda NTLP algorithm regardless of Xpert access. However, if CXR was unavailable, specificity was lower in WHO TDA-B compared to Uganda NTLP algorithm, across reference standards.

**Table 4:** Comparison of 2017 Uganda national childhood TB diagnosis algorithm (NTLP) against 2022 WHO childhood TB treatment decision algorithms among children <10 years old with and without Xpert testing or Chest X-ray imaging.

|  | NTLP |  | WHO Algorithm A/B<sup>1</sup> |  |  |  |
| --- | --- | --- | --- | --- | --- | --- |
| Scenario | Sensitivity %, (95%CI) | Specificity %, (95%CI) | Sensitivity %, (95%CI) | Specificity %, (95%CI) | Difference in Sensitivity (NTLP vs. WHO) % (95% CI), p-value | Difference in Specificity (NTLP vs. WHO) % (95% CI), p-value |
| Confirmed TB Vs. Unlikely TB |  |  |  |  |  |  |
| Xpert + CXR | 97.6 (95.9, 99.3) | 25.7(20.7, 30.7) | 96.3 (94.2, 98.5) | 32.2 (26.9, 37.5) | 1.3 (-2.3, 4.5), 1.00 | -6.8 (-11.9, -1.7), 0.01 |
| Xpert | 97.6 (95.9, 99.3) | 28.1 (22.9, 33.2) | 97.5 (95.8, 99.3) | 15.4 (11.3, 19.5) | 0 (-1.2, 1.2), 1.00 | 12.1 (5.3, 19.0), <0.01 |
| CXR | 93.7 (90.9, 96.5) | 25.7 (20.6, 30.7) | 96.3 (94.2, 98.5) | 32.2 (26.9, 37.5) | -2.5 (-3.7, 8.8), 0.62 | -6.8 (-11.9, -1.7), <0.01 |
| None | 91.2 (88.0, 94.5) | 28.1 (22.9, 33.2) | 97.5 (95.8, 99.3) | 15.4 (11.3, 19.5) | -6.4 (-0.3, 13.1), 0.06 | 12.1 (5.3, 19.0), <0.01 |
| Composite reference standard (CRS) |  |  |  |  |  |  |
| Xpert + CXR | 90.6 (88.3, 92.9) | 25.7 (22.2, 29.1) | 89.3 (86.8, 91.7) | 32.2 (28.5, 35.9) | 1.0 (-1.9, 4.0), 0.58 | -6.8 (-11.9, -1.7), <0.01 |
| Xpert | 88.9 (86.5, 91.4) | 28.1 (24.5, 31.6) | 94.2 (92.4, 96.1) | 15.4 (12.5, 18.2) | -5.6 (2.1, 9.2), <0.01 | 12.1 (5.3, 19.0), <0.01 |
| CXR | 89.8 (87.4, 92.2) | 25.7 (22.2, 29.1) | 89.3 (86.8, 91.7) | 32.2 (28.5, 35.9) | 0 (-2.9, 3.4), 1.00 | -6.8 (-11.9, -1.7), <0.01 |
| None | 87.5 (84.9, 90.2) | 28.1 (24.5, 31.6) | 94.2 (92.4, 96.1) | 15.4 (12.5, 18.2) | -7.0 (-10.7, -3.2), <0.01 | 12.1 (5.3, 19.0), <0.01 |
| Microbiologic reference standard (MRS) |  |  |  |  |  |  |
| Xpert + CXR | 97.6 (96.4, 98.8) | 16.8 (13.9, 19.8) | 96.3 (94.9, 97.8) | 20.4 (17.2, 23.5) | 1.2 (-2.3, 4.7), 1.00 | -3.3 (-0.3, 6.2), 0.02 |
| Xpert | 97.6 (96.4, 98.8) | 19.1 (16.0, 22.2) | 97.5 (96.3, 98.8) | 10.1 (7.7, 12.4) | 0 (-1.2, 1.2), 1.00 | 9.1 (5.4, 12.9), <0.01 |
| CXR | 93.7 (91.8, 95.6) | 16.8 (13.9, 19.8) | 96.3 (94.9, 97.8) | 20.4 (17.2, 23.5) | -2.5 (-3.7, 8.8), 0.62 | -3.3 (-0.3, 6.2), 0.02 |
| None | 91.2 (89.0, 93.4) | 19.1 (16.0, 22.2) | 97.5 (96.3, 98.8) | 10.1 (7.7, 12.4) | -6.4 (-0.3, 13.1), 0.06 | 9.1 (5.4, 12.9), <0.01 |

Xpert: Xpert MTB/RIF Ultra; CXR: Chest x-ray
1. Algorithm A used in scenarios with CXR, Algorithm B in scenarios without CXR

## DISCUSSION

In this study, we sought to determine the accuracy of the Uganda NTLP algorithm and also compare its accuracy to the WHO TDAs. We found that the Uganda NTLP algorithm had a high sensitivity but low specificity, under both microbiologic and composite reference standards, and the accuracy remained similar (high sensitivity and low specificity) in key risk groups (children < 5 years, CHIV, or SAM). Both the WHO TDA and the Uganda NTLP algorithms had similarly high sensitivity, but specificity was higher for the WHO TDA-A in settings with CXR available, while the Uganda NTLP algorithm had higher specificity in settings without molecular testing and/or CXR. Our findings point to the need for tools to augment the specificity of these algorithms, especially if molecular testing or CXR is not available.

The high sensitivity of the Uganda NTLP algorithm supports the goal of reducing missed TB cases, and previous work has shown that this algorithm increased TB notification among children (<15 years of age) in two Ugandan districts by 41% over an 18-month period.^[11]^ However, our study raises concern that a large proportion of children are overtreated for TB. As children in the general population are already screened first for symptoms or risk factors prior to utilizing the Uganda NTLP algorithm, greater specificity is needed. For the CRS, we found a small reduction in sensitivity when children with Unconfirmed TB were included, as expected given this is a heterogenous group and may include children without true TB disease. Our results were in contrast to an individual participant data (IPD) meta-analysis that showed the Uganda NTLP algorithm to have a lower sensitivity of 67% and higher specificity of 39%,^[12]^ though there was wide heterogeneity across studies, with a range of 33-97% sensitivity and 15-85% specificity. However, across reference standards, the sensitivity remained high and specificity was low, with implications for overtreatment of children. This was also true in subgroup analysis looking at CHIV, SAM, or below 5 years old. It may be appropriate to over-treat these higher risk groups, but further work is needed to understand the potential harms including side effects or missed alternative diagnoses.

The accuracy of the WHO TDAs was consistent with the IPD meta-analysis that found the pooled sensitivity and specificity at 80% and 30% respectively.^[12]^ Our higher sensitivity may be explained by the fact that children in our study were first screened using a more sensitive ICF, as similarly seen among adults in Kampala where there was a 6% increase in TB case notifications as a result of ICF implementation.^[13]^ Comparing the Uganda NTLP algorithm and WHO TDAs, the sensitivity remained high regardless of Xpert, reflecting that these have been developed in particular for empiric treatment when microbiological testing is not available or frequently negative, given paucibacillary disease in children. The Uganda NTLP algorithm’s accuracy remained stable with or without CXR, unlike the WHO TDA’s specificity that dropped in scenarios where CXR was unavailable. A possible reason for this is how each algorithm is scored. For the Uganda NTLP algorithm, the presence or absence of specific signs or symptoms are equally weighted, and so more children may be diagnosed with TB for example regardless of CXR findings. In contrast, the WHO TDAs assign points that differentially weight signs and symptoms, including specific CXR findings. Moreover, the assigned points are different whether CXR is present or not, and the internal validation data showed that the model with CXR had greater specificity.^[12]^ From an implementation perspective, the WHO TDAs and algorithms with a similar approach may be most valuable in settings with both Xpert and CXR available, but those are facilities where providers may have more experience with diagnosing TB. Although the Uganda NTLP algorithm had lower specificity, this was higher than WHO algorithm B and it remained stable with or without CXR; it therefore may be preferable in lower-level health facilities which may not have molecular or radiological testing facilities. Policy makers will need to balance the benefits of detecting more cases with the costs of over-treatment and expanding access to molecular testing with CXR to improve specificity.

Our study adds much needed data on the accuracy of the Uganda NTLP algorithm and its comparison to the WHO TDAs. We further present the accuracy results based on availability of CXR and Xpert to guide how these algorithms may perform in different types of settings. However, our study had limitations. The CRS is challenging because Unconfirmed TB can be greatly influenced by provider treatment initiation, which in turn may have been informed by the NTLP algorithm given it is implemented at the facility. Therefore, we will have high incorporation bias as the CRS includes both Xpert, CXR and clinical decision to treat. Another limitation is that we looked at urban children with presumptive TB and therefore our results may not generalize to rural or migratory populations. Additionally, the assessment was done by study staff focused on the evaluation of TB and all children provided respiratory samples, which could reduce generalizability. Also, our analysis was restricted to only children who fulfilled the ICF criteria; however, only eight children were excluded as a result of this requirement.

In summary, our study shows that if implemented in its current form, the Uganda NTLP algorithm and WHO TDAs have high sensitivity to capture most cases of childhood TB, but may result in overdiagnosis and overtreatment of children without TB. Our study further highlights a potential limitation of implementing the WHO TDAs over the Uganda NTLP algorithm in primary care clinics without access to CXR or molecular testing. As new TB diagnostics emerge, it will be important to integrate them into TDAs to assess incremental changes in accuracy.

## Data Availability

Data set uploaded as supplementary material

## ACKNOWLEDGMENT

We thank the patients, families, and staff at Mulago National Referral Hospital, Kampala Capital City Authority Clinics, and Infectious Diseases Institute (IDI) clinics. The study was supported by the National Institutes of Health (R01HL139717 and U01AI152087 to AC, and R01HL169449 and K23HL153581 to DJ). The content is solely the responsibility of the authors and does not necessarily represent the official views of the National Institutes of Health.

